# Surgical activity in England and Wales during the COVID-19 pandemic: a nationwide observational cohort study

**DOI:** 10.1101/2021.02.27.21252593

**Authors:** T D Dobbs, J A G Gibson, A J Fowler, T E Abbott, T Shahid, F Torabi, R Griffiths, R A Lyons, R M Pearse, I S Whitaker

**Affiliations:** Reconstructive Surgery & Regenerative Medicine Research Group, Institute of Life Sciences, Swansea University Medical School, Swansea, UK; Welsh Centre for Burns and Plastics, Morriston Hospital, Swansea, UK; William Harvey Research Institute, Queen Mary University of London, UK; Population Data Science, Swansea University Medical School Swansea, Wales, UK

**Author notes:** Correspondence to: Thomas Dobbs, Reconstructive Surgery & Regenerative Medicine Research Group, Institute of Life Sciences, Swansea University Medical School, Swansea, SA2 8PP UK. Joint first authors (T D Dobbs, J A G Gibson, A J Fowler, T E Abbott).

**Keywords:** Surgery, Anaesthesia, COVID-19, Public Policy

## Abstract

**Objectives:** To report the volume of surgical activity and the number of cancelled surgical procedures during the COVID-19 pandemic.

**Design and setting:** Analysis of electronic health record data from the National Health Service (NHS) in England and Wales.

**Methods:** We used hospital episode statistics for all adult patients undergoing surgery between 1^st^ January 2020 and 31^st^ December 2020. We identified surgical procedures using a previously published list of procedure codes. Procedures were stratified by urgency of surgery as defined by NHS England. We calculated the deficit of surgical activity by comparing the expected number of procedures from the years 2016-2019 with the actual number of procedures in 2020. We estimated the cumulative number of cancelled procedures by 31^st^ December 2021 according patterns of activity in 2020.

**Results:** The total number of surgical procedures carried out in England and Wales in 2020 was 3,102,674 compared to the predicted number of 4,671,338. This represents a 33.6% reduction in the national volume of surgical activity. There were 763,730 emergency surgical procedures (13.4% reduction), compared to 2,338,944 elective surgical procedures (38.6% reduction). The cumulative number of cancelled or postponed procedures was 1,568,664. We estimate that this will increase to 2,358,420 by 31^st^ December 2021.

**Conclusions:** The volume of surgical activity in England and Wales was reduced by 33.6% in 2020, resulting in over 1,568,664 cancelled operations. This deficit will continue to grow in 2021.

**Summary boxes:** *What is already known on this topic:*

- The COVID-19 pandemic necessitated a rapid change in the provision of care, including the suspension of a large proportion of surgical activity
- Surgical activity has yet to return to normal and has been further impacted by subsequent waves of the pandemic
- This will lead to a large backlog of cases

*What this study adds:*

- 3,102,674 surgical procedures were performed in England and Wales during 2020, a 33.6% reduction on the expected yearly surgical activity
- Over 1.5 million procedures were not performed, with this deficit likely to continue to grow to 2.3 million by the end of 2021
- This deficit is the equivalent of more than 6 months of pre-pandemic surgical activity, requiring a monumental financial and logistic challenge to manage

## Introduction

The COVID-19 pandemic has fundamentally changed the delivery of healthcare worldwide. In many health systems, resources have been reallocated to the care of patients with acute SARS-CoV-2 infection and diverted away from routine healthcare services including cancer care and chronic disease management.^1,2^ Critical care capacity was increased, staff and equipment were redeployed and, in some cases, new hospitals were built.^3-5^ In England, the National Health Service (NHS) postponed all non-urgent surgery from 15^th^ April 2020 to support the response to the first wave of the pandemic, with repetition of these delays from December 2020 in response to the second wave.^6-8^ This has had an enormous, but as yet uncharacterised, impact on the provision of surgery, resulting in the cancellation of a large portion of elective surgery and delayed urgent surgery for almost an entire year.

In high-income countries, surgical services represent a large portion of healthcare activity.^9^ In the NHS, surgery accounts for over 5 million hospital admissions every year.^10,11^ However, the exact number of surgical procedures that have been cancelled due to COVID-19 is unknown. During 2020, there was a phased reintroduction of urgent and elective surgery following the first lockdown. However, this was complicated by increased staff sickness, reduced operating room capacity and lower throughput due to enhanced infection control policies. In the meantime, the waiting list for surgical procedures has continued to grow.^12^ In a previous study, we estimated that over two million surgical procedures would have been cancelled in the NHS in England due to the first wave of the pandemic.^13^ However, this was modelled on previous years data and anecdotally observed falls in activity, as well as not accounting for a second wave of COVID-19 cases and further associated reductions in surgical activity. Therefore, the true impact of COVID-19 on national surgical activity is still unknown. It is crucial that we understand the total number of cancelled surgical procedures, the size of waiting list and the priority of these cases, so that healthcare leaders and policy makers are able to plan the reintroduction of urgent and elective surgery.

Here, we report the results of a planned analysis of hospital episode data from the NHS in England and Wales. We describe the true national volume of surgical activity that occurred during the first wave of the COVID-19 pandemic and the initial recovery period, and also estimate the likely number of cancelled procedures until the end of 2021.

## Methods

### Study design and setting

*P*opulation-based epidemiological study describing all hospital admissions for a surgical procedure in the National Health Service (NHS) in England and Wales between 1^st^ January 2020 and 31^st^ December 2020. We used data from 1^st^ January 2016 until 31^st^ December 2019 as a historical comparator period.

### Data sources

We used Hospital Episode Statistics for Admitted Patient Care (HES APC) and Patient Episode Data for Wales (PEDW), which describe every episode of hospital care in the NHS in England and Wales, respectively. The databases include demographic information (age, sex, ethnicity), process information (dates of admission, start of episode, end of episode, reason for admission and patient class), and procedural information (recorded using Office of Population Censuses and Surveys classification of interventions and procedures version 4.7 [OPCS-4.7] codes and associated dates).^14,15^

### Study population

All patients undergoing surgical procedures in England and Wales in the study period were included. Surgical procedures were defined using a previously published definition of surgery identified using three-character OPCS-4.7 codes (Appendix A).^10,11^

### Outcomes

The primary outcome was the number of hospital admissions with an associated surgical procedure.

### Data processing

Duplicate episodes were removed and procedures were restricted to primary procedures within each admission episode. This included finished consultant episodes only, which excludes hospital episodes where a patient remains in hospital, or where data has not yet been submitted to NHS England/NHS Wales. Data were extracted from core data tables in the Data Access Environment (England) and the SAIL Databank (Wales). We selected all hospital episodes associated with defined OPCS-4.7 codes.^10,11^. We categorised each procedure according to urgency of the surgical procedure using four classes defined by NHS England (Appendix B): Emergency (required within 72 hours); Urgent (can be delayed for up to four weeks); Semi-urgent (can be delayed for up to three months); Elective (can be delayed for more than three months).^6^ The OPCS 4.7 contributing to these classes were derived from average waiting times reported between 2014 to 2019, as previously described.^16^ We aggregated the data by anatomical location, month, and year of the procedure (Appendix A).10,13

### Statistical analysis

#### Volume of procedures

Descriptive statistics were used to characterise the volume of procedures performed during the historical comparative period (2016-2019) and during 2020.

#### Estimation of the deficit of surgical volume

The expected monthly frequency of surgical activity from 1^st^ January 2020 to 31^st^ December 2020 was estimated using the surgical activity during the three-years prior to the study period (1^st^ January 2016 to 31^st^ December 2019). A time series model was used to independently forecast the total monthly number of procedures and the monthly number of procedures for each of the aforementioned surgical classifications.

#### Modelling analysis

We calculated the deficit of surgical procedures on 31^st^ December 2020 with a 95% confidence interval, based on the difference between the expected and actual number of cases that took place in the year to 31^st^ December 2020. We used a linear regression model to determine the rate of surgical recovery between the first two waves of the pandemic (April to October 2020). We assumed that surgical services would remain disrupted until the end of March 2021 and extrapolated growth from that baseline using the linear model to determine when surgery would reach normal capacity. All data were extracted using SQL and analysed using IBM SPSS Statistics for Windows (IBM Corp. Released 2017. Version 25.0. Armonk, NY: IBM Corp). Graphs were made using *ggplot2* package in R (R version 4.0.1, R Core Team, core team; Vienna).

#### Research ethics approval

This analysis of routinely collected, pseudonymised data was approved by the Health Research Authority (20/HRA/3121). Analysis of NHS Wales data was approved by SAIL independent Information Governance Review Panel (IGRP) project number 0911. Access to NHS England data was approved by the NHS Digital Independent Group Advising on the Release of Data (DARS-NIC-375669-J7M7F).

## Results

We identified 3,102,674 admissions for a surgical procedure between 1^st^ January 2020 and 31^st^ December 2020 (Appendix C). Of these, 2,981,161 (96%) were in England and 121,513 were in Wales (Appendix D). The median age was 58 (IQR: 38 to 73) years and 54% of the cohort were female.

### Comparison with the average annual activity 2016-2019

During the historical period (1^st^ January 2016 to 31^st^ December 2019), there were a median of 4,685,106 admissions for surgery per year (IQR 4,4,640,122 – 4,731,338). There were a median of 392,720.5 admissions per month (IQR 376,400.3 -406,084.3). The number of procedures expected to have been performed during 2020 was 4,671,338 (95% CI: 4,218,740 to 5,123,932). During 2020, the lowest number of surgical admissions were observed in the month of April with 104,063 (compared to the expected number of 381,153 representing a 72.7% reduction in surgical activity during this month). The number of admissions during the pandemic period, compared to the expected number of admissions is presented in Table 1 and Figure 1.

**Table 1.** Observed number of procedures vs expected number of procedures.

|  | Observed<br>, N | Class 1<br>Percentage of<br>expected (95%CI) | Observed, N | Class 2<br>Percentage of<br>expected (95%CI) | Observed,<br>N | Class 3<br>Percentage of<br>expected (95%CI) | Observed, N | Class 4<br>Percentage of expected<br>(95%CI) |
| --- | --- | --- | --- | --- | --- | --- | --- | --- |
| Jan-20 | 72,528 | 99.8 (92.6-108.2) | 35,067 | 106 (95.4-119.2) | 216,455 | 101.5 (91.6-113.7) | 75,195 | 98.0 (84.0-117.5) |
| Feb-20 | 67,141 | 92.4 (85.7-100.1) | 31,839 | 96.2 (86.6-108.2) | 201,663 | 97.7 (87.9-110.0) | 72,343 | 90.3 (77.9-107.3) |
| Mar-20 | 62,355 | 85.8 (79.6-93.0) | 29,629 | 89.5 (80.5-100.7) | 149,742 | 69.5 (62.8-77.8) | 44,496 | 57.3 (49.7-69.5) |
| Apr-20 | 48,798 | 67.1 (62.3-72.8) | 16,525 | 49.9 (44.9-56.1) | 37,120 | 18.4 (16.5-20.7) | 1,620 | 2.3 (1.9-2.7) |
| May-20 | 59,597 | 82.0 (76.1-88.9) | 18,012 | 54.4 (48.9-61.2) | 47,274 | 22.9 (20.6-25.8) | 3,542 | 4.5 (3.8-5.5) |
| Jun-20 | 64,178 | 88.3 (82.0-95.7) | 21,772 | 65.7 (59.1-73.9) | 77,975 | 38.2 (34.4-43.1) | 11,033 | 14.1 (12.0-17.3) |
| Jul-20 | 70,633 | 97.2 (90.2-105.3) | 24,988 | 75.4 (67.8-84.8) | 113,141 | 53.2 (48.0-59.6) | 23,619 | 30.3 (25.6-36.9) |
| Aug-20 | 68,898 | 94.8 (88.0-102.8) | 23,183 | 69.9 (62.9-78.6) | 118,748 | 59.1 (53.0-66.8) | 31,785 | 40.7 (34.3-49.7) |
| Sep-20 | 68,942 | 94.9 (88.0-102.8) | 27,145 | 81.8 (73.6-92.0) | 155,569 | 74.9 (67.5-84.3) | 46,239 | 59.2 (50.1-72.3) |
| Oct-20 | 67,314 | 92.6 (86.0-100.4) | 27,933 | 84.1 (75.7-94.7) | 168,026 | 79.4 (71.6-89.1) | 53,518 | 68.6 (58.0-83.7) |
| Nov-20 | 62,125 | 85.5 (79.3-92.6) | 27,007 | 81.3 (73.2-91.5) | 164,060 | 79.4 (71.4-89.4) | 50,154 | 64.2 (54.4-78.5) |
| Dec-20 | 51,221 | 70.5 (65.4-76.2) | 24,313 | 73.2 (65.8-82.3) | 133,035 | 67.4 (60.4-76.3) | 35,179 | 45.1 (38.2-55.0) |

*Class 1, Emergency surgery with 72 hours; Class 2, Urgent surgery within 4 weeks; Class 3, Semi-urgent surgery within 3 months; Class 4, Elective surgery over 3 months*

**Figure 1.**
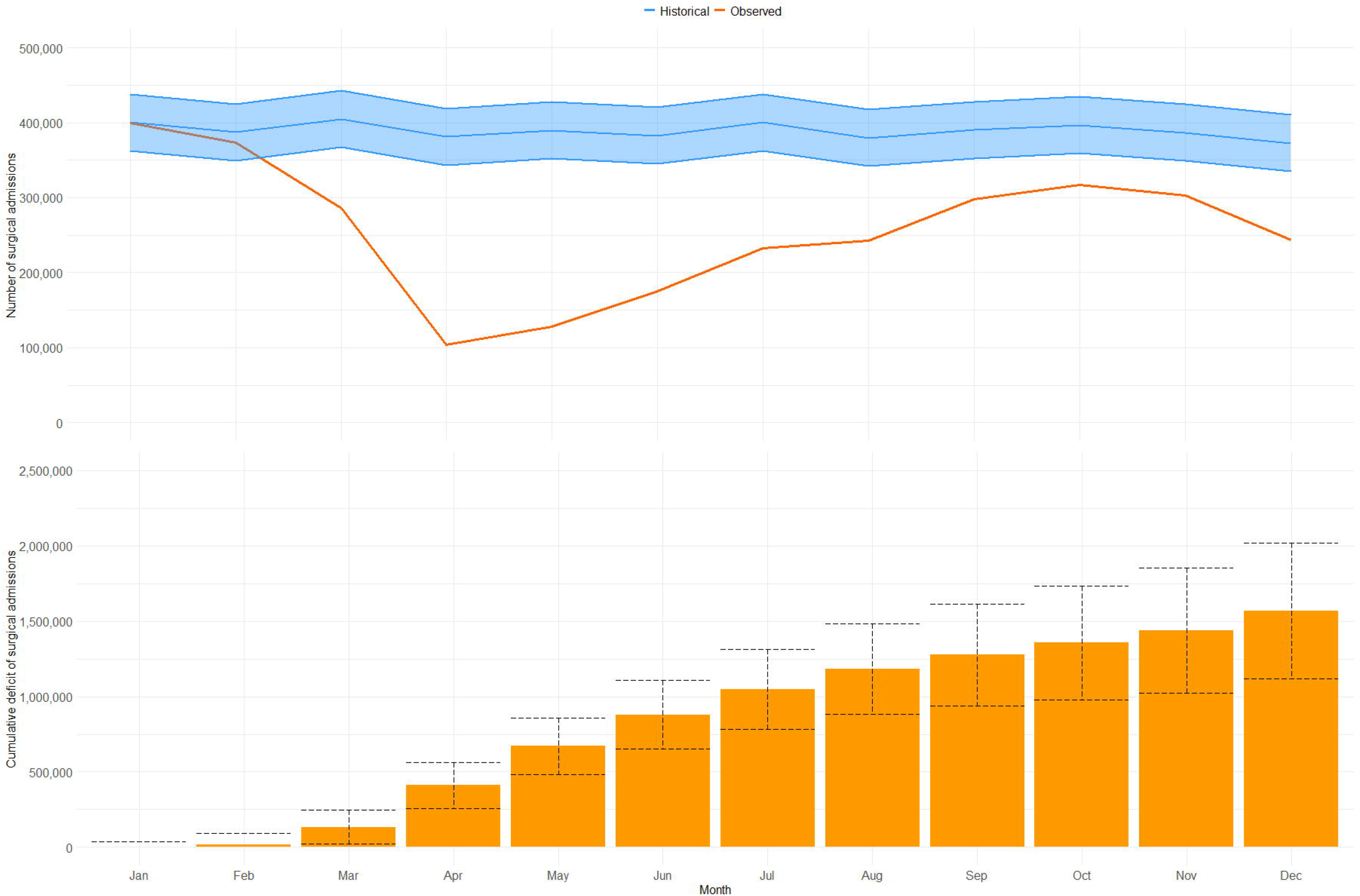
Top panel: Surgicwal activity during 2020 (red line) compared to expected surgical activity (blue line) based on the years 2016-2019 (with 95% CIs). Bottom panel: Cumulative deficit of surgical procedures throughout 2020 (95% CIs).

### Deficit of surgical activity

The total cumulative deficit of hospital admissions for surgical procedures on the 31^st^ December 2020 was 1,568,664 (95%CI: 1,116,066 to 2,021,258) procedures (Figure 1). There were 763,730 emergency surgical procedures (13.4% reduction), compared to 2,338,944 elective surgical procedures (38.6% reduction). The majority of this deficit are represented by semi-urgent surgical procedures in class 3 (904,761 of 1,568,664; 57.7%) and elective surgical procedures in class 4 (481,150 of 1,568,664 30.7%) (Table 2).

**Table 2.** Monthly and cumulative deficit of number of surgical procedures.

|  | Total |  | Class 1 |  | Class 2 |  | Class 3 |  | Class 4 |  |
| --- | --- | --- | --- | --- | --- | --- | --- | --- | --- | --- |
|  | Monthly deficit | Cumulative deficit | Monthly deficit | Cumulative deficit | Monthly deficit | Cumulative deficit | Monthly deficit | Cumulative deficit | Monthly deficit | Cumulative deficit |
| January | 863 | 863 | 150 | 150 | 0 | 0 | 0 | 0 | 1,550 | 1,550 |
| February | 14,055 | 14,918 | 5,537 | 5,687 | 1,257 | 1,257 | 4,773 | 4,773 | 7,811 | 9,361 |
| March | 118,492 | 133,410 | 10,323 | 16,010 | 3,480 | 4,737 | 65,692 | 70,465 | 32,291 | 41,652 |
| April | 277,090 | 410,500 | 23,880 | 39,890 | 16,597 | 21,334 | 165,060 | 235,525 | 70,055 | 111,707 |
| May | 261,176 | 671,676 | 13,081 | 52,971 | 15,123 | 36,457 | 159,167 | 394,692 | 74,522 | 186,229 |
| June | 207,880 | 879,556 | 8,500 | 61,471 | 11,376 | 47,833 | 125,974 | 520,666 | 67,031 | 253,260 |
| July | 167,730 | 1,047,286 | 2,045 | 63,516 | 8,173 | 56,006 | 99,650 | 620,316 | 54,445 | 307,705 |
| August | 137,072 | 1,184,358 | 3,780 | 67,296 | 9,990 | 65,996 | 82,126 | 702,442 | 46,279 | 353,984 |
| September | 92,131 | 1,276,489 | 3,736 | 71,032 | 6,041 | 72,037 | 51,997 | 754,439 | 31,825 | 385,809 |
| October | 79,742 | 1,356,231 | 5,364 | 76,396 | 5,266 | 77,303 | 43,510 | 797,949 | 24,546 | 410,355 |
| November | 83,499 | 1,439,730 | 10,553 | 86,949 | 6,205 | 83,508 | 42,533 | 840,482 | 27,910 | 438,265 |
| December | 128,934 | 1,568,664 | 21,457 | 108,406 | 8,912 | 92,420 | 64,279 | 904,761 | 42,885 | 481,150 |

*Class 1, Emergency surgery with 72 hours; Class 2, Urgent surgery within 4 weeks; Class 3, Semi-urgent surgery within 3 months; Class 4, Elective surgery over 3 months*

### Number of surgical admissions by urgency of procedure

In 2020 the majority of surgical activity were class 3 (1,582,808 of 3,102,674, 51.0%). The greatest reduction in activity compared to the historical cohort was seen in class 4 procedures (448,723 of 929,873 a 51.8% reduction). The number of admissions during 2020, compared to the expected number of admissions categorised by class is presented in Table 1 and Figure 2.

**Figure 2.**
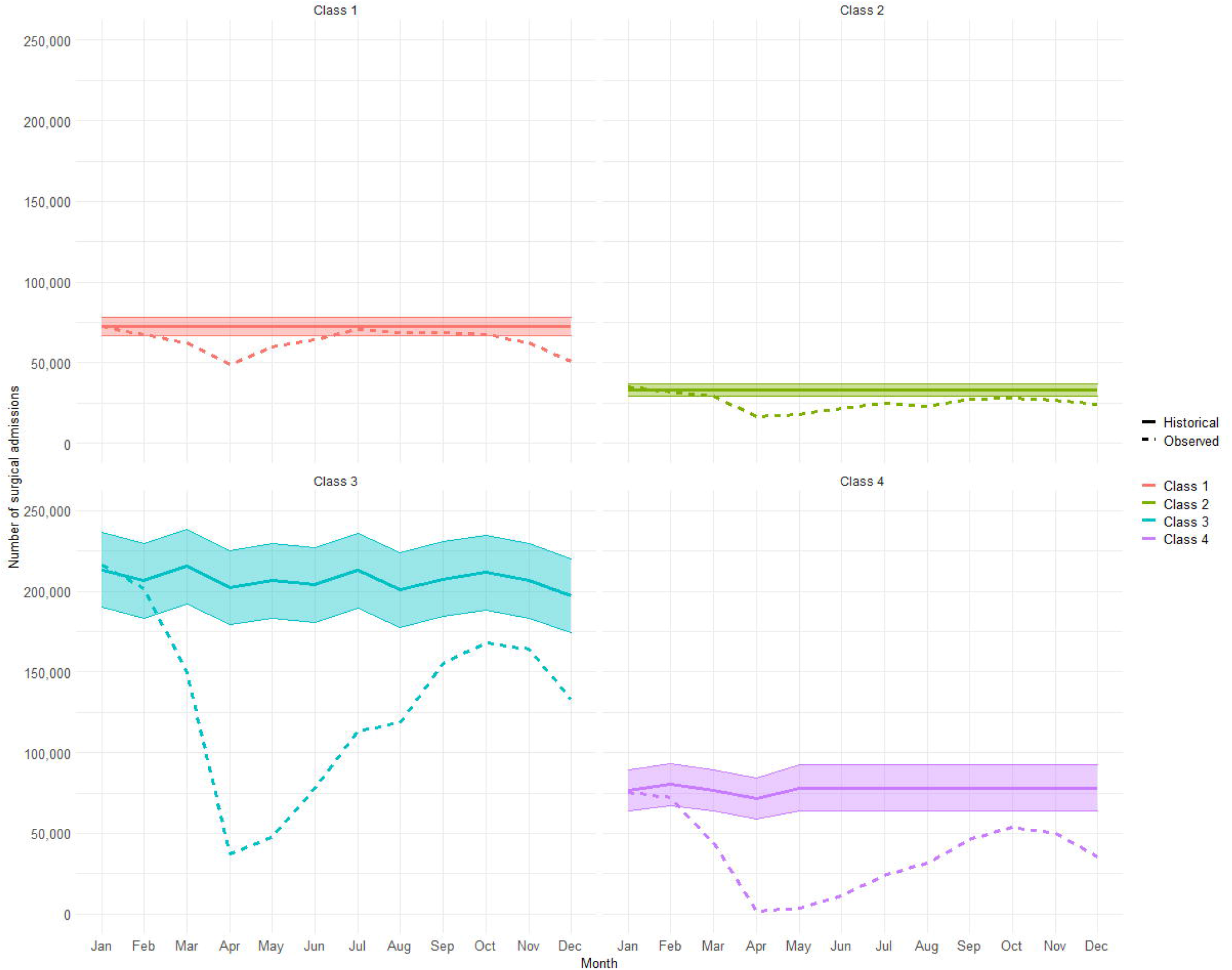
Surgical activity by class for 2020 (dotted line) compared to expected surgical activity and 95% CI (solid line and shading).

### Number of surgical admissions by specialty

The number of admissions for surgery during 2020, stratified by anatomical surgical site is provided in Appendix E. The surgical specialty with the biggest reduction in activity during 2020 compared to the historical comparator was Oral Surgery (48 %).

### Predicted volume of surgical activity in 2021

If the reintroduction of surgical activity in March 2021 occurs at a similar rate to that observed between the first and second waves of the pandemic (April to October 2020), then surgical care will reach pre-pandemic activity levels by August 2021 (Figure 3). We estimate a cumulative deficit of 2,384,200 (95% CI: 1,667,587 to 3,100,808) surgical procedures between 1^st^ January 2020 and 31^st^ December 2021 (Table 3). Based on the median monthly number of procedures performed during 2016-2019, this is equivalent to more than six months of pre-pandemic surgical activity.

**Table 3.** Modelled number of procedures, monthly deficit, and cumulative deficit in 2021. Data are counts of procedures with associated 95% confidence interval derived from a linear regression model.

| **2021:** | **Modelled number of procedures** | **Monthly deficit compared to historical period** | **Cumulative deficit (2021 only)** |
| --- | --- | --- | --- |
| **January** | 243,748 (243,748 to 243,748) | 156,360 (118,644 to 194,076) | 156,360 (118,644 to 194,076) |
| **February** | 243,748 (243,748 to 243,748) | 143,293 (105,577 to 181,010) | 299,653 (224,221 to 375,086) |
| **March** | 243,748 (243,748 to 243,748) | 160,966 (123,249 to 198,682) | 460,619 (347,470 to 573,768) |
| **April** | 243,726 (210,656 to 276,797) | 137,427 (99,710 to 175,143) | 598,046 (447,180 to 748,911) |
| **May** | 281,040 (246,677 to 315,402) | 108,561 (70,845 to 146,278) | 706,607 (518,025 to 895,189) |
| **June** | 318,353 (281,817 to 354,890) | 64,485 (26,769 to 102,201) | 771,092 (544,794 to 997,390) |
| **July** | 355,667 (316,220 to 395,114) | 44,444 (6,727 to 82,160) | 815,536 (551,521 to 1,079,550) |
| **August** | 379,686 (341,969 to 417,402) | 0 (0 to 0) | 815,536 (551,521 to 1,079,550) |
| **September** | 390,026 (352,309 to 427,742) | 0 (0 to 0) | 815,536 (551,521 to 1,079,550) |
| **October** | 396,533 (358,817 to 434,249) | 0 (0 to 0) | 815,536 (551,521 to 1,079,550)) |
| **November** | 386,845 (349,129 to 424,561) | 0 (0 to 0) | 815,536 (551,521 to 1,079,550) |
| **December** | 372,682 (334,965 to 410,398) | 0 (0 to 0) | 815,536 (551,521 to 1,079,550) |

**Figure 3.**
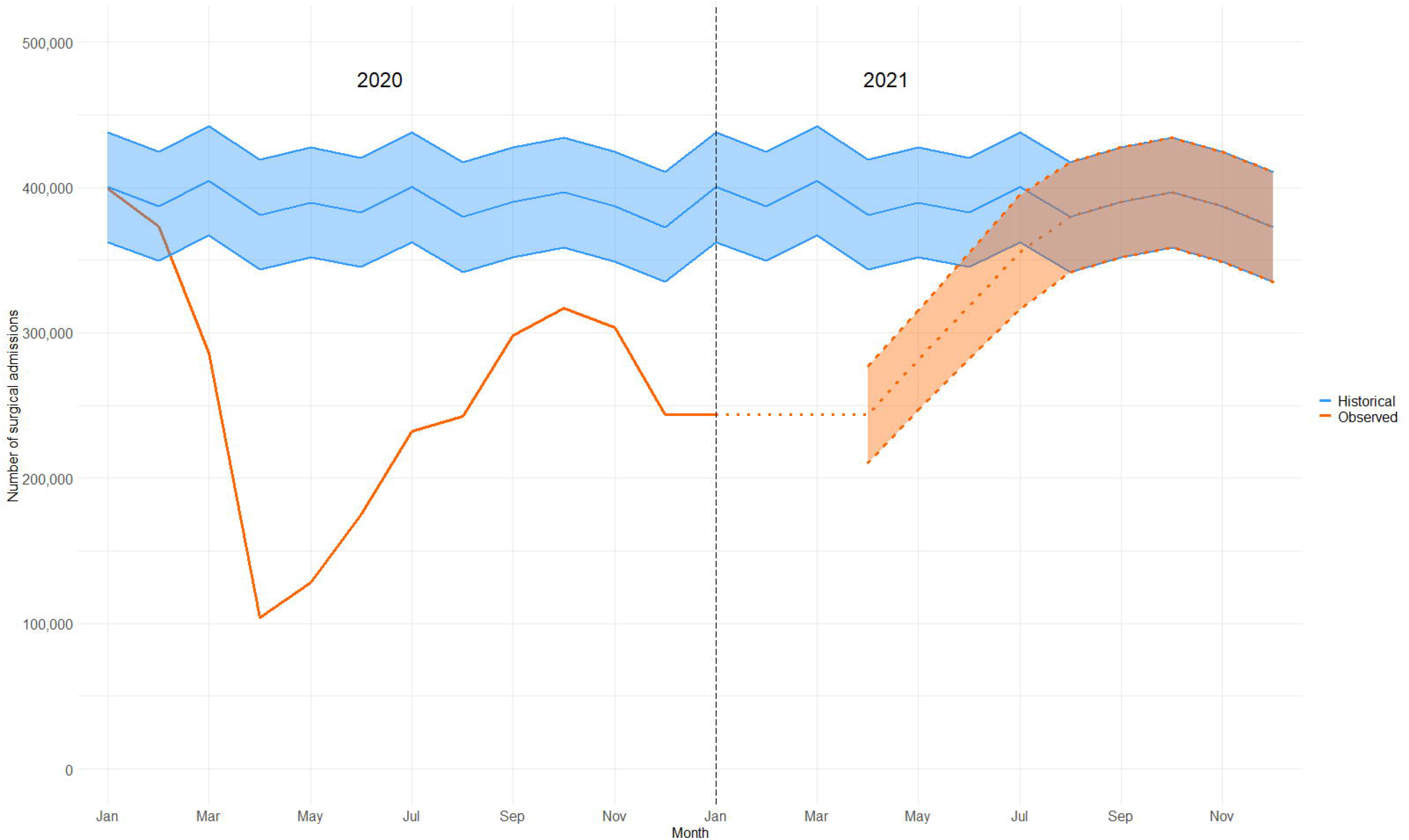
Predicted surgical activity for 2021 and return to pre-pandemic operative volume.

## Discussion

The principle finding of this national observational study is that the overall volume of surgical activity in England and Wales during 2020 is 33.6% lower than expected levels. This represents the cancellation or postponement of more than 1.5 million surgical procedures. The vast majority of the deficit of procedures is accounted for by semi-urgent and elective surgery (38.6%). However, there was a substantial reduction in the frequency of emergency surgery (13.4%), which has not yet returned to a pre-pandemic baseline. This may be due to a reduction in emergency surgical presentations, for example fewer injuries requiring surgery due to lockdowns; it may represent an inflation of emergency cases numbers before the pandemic (i.e. operations that were not true emergencies) or clinical management using non-surgical treatment (i.e. conservative or medical therapy). We predict that by the end of 2021 there will 2.4 million surgical procedures outstanding, representing more than six months of normal surgical activity.

Perioperative clinicians in the United Kingdom and worldwide will be familiar with changes to working patterns, reductions in operating theatre capacity and increasing waiting lists for surgery.^5,17^ Several research groups have estimated the volume of cancelled surgical procedures.^8,13,18^ However, this is the first study, to our knowledge, to describe the impact of the COVID-19 pandemic on the volume of surgical activity at the level of national health systems. Our data represent the true reduction in surgical workload that occurred during the first wave of the pandemic in England and Wales, and the subsequent (incomplete) recovery of surgical services. In the early phase of the pandemic there was a necessary trade off between care of a large volume patients with acute respiratory disease, many of whom required intensive care treatments, and continuation of services to treat surgical disease. This led to the almost complete cessation of all but the most emergent surgical treatment. The reintroduction of surgery after the first wave of the pandemic was complicated by several factors. First, concerns about the high-risk of perioperative mortality among surgical patients with concomitant SARS-CoV-2 infection and the postponement of surgery on clinical grounds. ^17,19^ Second, reduced availability of physical and human resources due to on-going care of patients with COVID-19, which limited the capacity of surgical services.^20^ Third, reduced throughput of surgery due to new infection control procedures to prevent nosocomial SARS-CoV-2 infection and to protect staff members. The consequences of the observed interruption to surgical treatment will be felt by millions of patients for many years to come. Delays in the diagnosis and the surgical management of cancer will undoubtably lead to an increase in cancer related mortality.^21^ For many patients waiting for semi-urgent and elective surgery, there is likely to be a worsening of their condition while waiting for surgery, which could make future treatment more difficult and less likely to succeed.^22^ Furthermore these patients are likely to suffer worsening of their physical and mental health while on the waiting list, placing additional burdens on primary and social care, reducing their productivity in the workplace.^23^ This represents a huge financial and human cost to society.

Dealing with the backlog of surgery will be a challenge to many health systems worldwide.^24^ We have previously demonstrated that SARS-CoV-2-free ‘green pathway’ operating is safe, with similar perioperative mortality among patients on ‘green pathways’ compared to before the pandemic.^19^ Consequently, surgery should not be delayed or prevented if safeguards are put in place. However, this will require significant re-organisation of surgical services and financial commitment from central government. Green pathway operating will need dedicated space and staff to ensure safety, with routine SARS-CoV-2 testing for patients and staff. With limited personnel and physical resources, it may take many years of concerted effort to clear the backlog of surgery due to COVID-19. In the rush to limit harm to surgical patients and restart elective surgery as soon as possible, staff welfare should not be neglected. A high proportion of healthcare workers have experienced burn out over the last year and adequate time for staff recovery should be incorporated into any plan to tackle the waiting list for surgery.^25^ Asking staff that are already tired to work harder for longer may not be a sustainable solution to this problem.

This study has several strengths. We included data from all patients undergoing surgery in England and Wales during the first wave of the COVID-19 pandemic, thus our results represent the true volume of surgery being performed. We include data from 2.5 million surgical patients, which represents one of the largest observational cohorts of surgical patients during the COVID-19 pandemic. A pre-pandemic cohort of more than 10 million patients was used to accurately identify the expected usual surgical activity within the NHS for comparison. Official routine data collection is also less open to bias, which can affect other data collection methods such as clinician surveys. Furthermore, this data will be generalisable to other high-income countries, where similar reductions in surgical activity have been reported.^26^ The pandemic cohort time period was chosen for a number of reasons. While the NHS response to COVID-19 began in early March 2020 there is evidence of community transmission within the UK prior to this.^27^ Furthermore, it is important that the winter months (January/February/March) were included in this analysis as operating volume is traditionally reduced due to other illnesses at this time of year. Our cohort cut off of the 31^st^ December 2020 allowed us to analyse the first wave and subsequent recovery, leading into the second wave of the pandemic, whilst still allowing for data to feed through to national databases and be analysed in a timely manner. This study also has limitations. As with any population-based study using routinely collected data there is the risk of incomplete or missing data, although from previous work this is likely to be negligible in this setting. Furthermore, the second peak of the pandemic did not occur until February 2021 in the UK, so it is likely that surgical activity continued to decline after December. We deliberately took a conservative approach in our modelling by assuming December was the nadir of surgical activity after April 2020.

## Conclusion

This study is the first to provide operational detail on surgical activity during the COVID-19 pandemic for an entire health care system. The volume of surgical activity in England and Wales in 2020 was 33.6% lower than historical data. Over 1.5 million surgical procedures have been cancelled as a result of COVID-19 and we predict that the deficit will rise to 2.4 million by the end of 2021. It is imperative that surgical patients are not the forgotten casualties of the pandemic. Further work must aim to identify ways to improve utilisation of all available capacity for surgical activity, with commitments by governments to make financing this work a priority.

## Data Availability

The data used in this study are derived from two data sources. It is not possible to share the raw patient-level data provided by NHS Digital describing NHS patients in England. Regarding data from NHS patients in Wales, the data used are available in the SAIL Databank at Swansea University, Swansea, UK, but as restrictions apply they are not publicly available. All proposals to use SAIL data are subject to review by an independent Information Governance Review Panel (IGRP). Before any data can be accessed, approval must be given by the IGRP. The IGRP gives careful consideration to each project to ensure proper and appropriate use of SAIL data. When access has been granted, it is gained through a privacy protecting safe haven and remote access system referred to as the SAIL Gateway. SAIL has established an application process to be followed by anyone who would like to access data via SAIL at https://www.saildatabank.com/application-process.

## Contributors

TA, TD, AF, RP were responsible for study design. JG, AF, FT, RG and RAL were responsible for data collection. JG and AF were responsible for data analysis. TD, TA, JG and AF were responsible for data interpretation. TD and TA wrote the first draft of the manuscript. All authors revised the manuscript for important intellectual content and approved the final version. AJF and JG had full access to the data and act as guarantors. The corresponding author attests that all listed authors meet the authorship criteria and that no others meeting the criteria have been omitted.

## Transparency declaration

TD affirms that the manuscript is an honest, accurate, and transparent account of the study being reported; that no important aspects of the study have been omitted; and that any discrepancies from the study as planned have been explained.

## Acknowledgements

This work uses data provided by patients and collected by the NHS as part of their care and support. We would like to acknowledge all data providers who make anonymised data available for research and the collaborative partnership that enabled acquisition and access to the de-identified data, which led to this output. The collaboration was led by the Swansea University Health Data Research UK team under the direction of the Welsh Government Technical Advisory Cell (TAC) and includes the following groups and organisations: the Secure Anonymised Information Linkage (SAIL) Databank, Administrative Data Research (ADR) Wales, NHS Wales Informatics Service (NWIS), Public Health Wales, NHS Shared Services and the Welsh Ambulance Service Trust (WAST). All research conducted has been completed under the permission and approval of the SAIL independent Information Governance Review Panel (IGRP) project number 0911.

## Patient and public involvement

A patient representative was consulted in the design of this study.

## Dissemination declaration

The results of this study have been shared with representatives from NHS England and NHS Wales prior to publication, in order to inform patient care.

## Licence

The Corresponding Author has the right to grant on behalf of all authors and does grant on behalf of all authors, a worldwide licence (http://www.bmj.com/sites/default/files/BMJ%20Author%20Licence%20March%202013.doc) to the Publishers and its licensees in perpetuity, in all forms, formats and media (whether known now or created in the future), to i) publish, reproduce, distribute, display and store the Contribution, ii) translate the Contribution into other languages, create adaptations, reprints, include within collections and create summaries, extracts and/or, abstracts of the Contribution and convert or allow conversion into any format including without limitation audio, iii) create any other derivative work(s) based in whole or part on the on the Contribution, iv) to exploit all subsidiary rights to exploit all subsidiary rights that currently exist or as may exist in the future in the Contribution, v) the inclusion of electronic links from the Contribution to third party material where-ever it may be located; and, vi) licence any third party to do any or all of the above. All research articles will be made available on an open access basis (with authors being asked to pay an open access fee— see http://www.bmj.com/about-bmj/resources-authors/forms-policies-and-checklists/copyright-open-access-and-permission-reuse). The terms of such open access shall be governed by a <u>Creative Commons</u> licence—details as to which Creative Commons licence will apply to the research article are set out in our worldwide licence referred to above.

Appendix A – Previous published definition of surgery using three-character Office of Population Censuses and Surveys classification of interventions and procedures version 4.7 (OPCS-4.7) codes.

Appendix B – Four class definition of the urgency of surgery as published by NHS England.

Appendix C – Flow diagram demonstrating number of patients identified in study cohort.

Appendix D - Surgical activity in 2020 classified by category of surgery for England and Wales.

Appendix D – Surgical activity in 2020 classified by anatomical site.

