## Appendices for "Surgical activity in England and Wales during the COVID-19 pandemic: a nationwide observational cohort study"

### SUPPLEMENTARY FILE

### Contents


| Item | Description | Page(s) |
|---|---|---|
| **Appendix A** | Previous published definition of surgery using three-character Office of Population Censuses and Surveys classification of interventions and procedures version 4.7 (OPCS-4.7) codes | **3-4** |
| **Appendix B** | OPCS 4.7 codes defining different elective procedure classes, stratified by anatomical grouping | **5-7** |
| **Appendix C** | Flow diagram demonstrating number of patients identified in study cohort | **8** |
| **Appendix D** | Surgical activity in 2020 classified by category of surgery for England and Wales | **9-10** |
| **Appendix E** | Surgical activity in 2020 classified by anatomical site | **11-14** |

| **Anatomical Location** | **OPCS 4.7 Codes** |
|---|---|
| Neuro | A01,A02,A03,A04,A05,A06,A07,A08,A09,A10,A11,A12,A13,A14,A16,A17,A18,A20,A22,A24,A25,A26,A27,A28,A29,A30,A31,A32,A33,A34,A36,A38,A39,A40,A41,A42,A43,A44,A45,A47,A48,A49,A51,A57,A59,A60,A61,A62,A63,A64,A65,A66,A67,A68,A69,A70,A73,A75,A76,A77,A78,A79,A81,A84,A82 |
| Endocrine | B01,B02,B04,B06,B08,B09,B10,B12,B14,B16,B17,B18,B20,B22,B23,B25 |
| Breast | B27,B28,B29,B30,B31,B33,B34,B35,B36,B37,B38,B39,B40 |
| Ocular | C01,C02,C03,C05,C06,C08,C09,C10,C11,C12,C13,C14,C15,C16,C17,C18,C19,C20,C22,C23,C24,C25,C26,C27,C29,C31,C32,C33,C34,C35,C37,C39,C40,C41,C43,C44,C45,C46,C47,C49,C51,C52,C53,C54,C55,C57,C59,C60,C61,C62,C64,C65,C66,C67,C69,C71,C72,C73,C74,C75,C77,C79,C80,C81,C82,C83,C84,C85,C86,C88,C89 |
| Ear | D01,D02,D03,D04,D06,D08,D10,D12,D13,D14,D15,D16,D17,D19,D20,D22,D23,D24,D26,D28 |
| Nasal | E01,E02,E03,E04,E05,E07,E08,E09,E10,E11,E12,E13,E14,E15,E16,E17,E64,E66 |
| Pharynx | E19,E20,E21,E23,E24,E27,E28,E29,E30,E31,E33,E34,E35,E38,F34 |
| Thoracic | E39,E40,E41,E42,E43,E44,E46,E47,E48,E50,E52,E53,E54,E55,E57,E59,E61,E62,E63,T01,T02,T03,T05,T07,T08,T09,T10,T11,T12,T13,T14,E67 |
| Oral | F01,F02,F03,F04,F05,F06,F09,F11,F18,F22,F23,F24,F26,F28,F29,F30,F32,F36,F38,F39,F40,F42,F44,F45,F46,F48,F50,F51,F52,F53,F58 |
| Upper GI | G01,G02,G03,G04,G05,G06,G07,G08,G09,G10,G11,G13,G14,G17,G21,G23,G24,G25,G26,G27,G28,G29,G30,G31,G32,G33,G34,G35,G36,G38,G40,G41,G48,G49,G50,G51,G52,G53,G57,G58,G59,G60,G61,G63,G67,G68,G69,G70,G71,G72,G73,G74,G75,G76,G78,G82,T15,T16,T17,T37,G12,G20 |
| Lower GI | H01,H02,H03,H04,H05,H06,H07,H08,H09,H10,H11,H12,H13,H14,H15,H16,H17,H19,H29,H30,H32,H33,H34,H35,H36,H40,H41,H42,H44,H46,H47,H48,H49,H50,H51,H52,H53,H54,H55,H56,H57,H58,H59,H60,H62,H66,T19,T20,T21,T22,T23,T24,T25,T26,T27,T28,T29,T30,T31,T33,T34,T36,T38,T39,T41,T42,T43,T45,T48,T51,T97,T98,X14,H37,T32 |
| HPB | J01,J02,J03,J04,J05,J06,J07,J08,J10,J11,J12,J13,J15,J16,J18,J19,J20,J21,J23,J24,J25,J26,J27,J28,J29,J30,J31,J32,J33,J34,J35,J36,J37,J49,J52,J54,J55,J56,J57,J58,J59,J60,J61,J62,J63,J65,J68,J69,J70,J72,J73,J77 |
| Cardiac | K01,K02,K04,K05,K06,K07,K08,K09,K10,K11,K12,K13,K14,K15,K16,K17,K18,K19,K20,K22,K23,K24,K25,K26,K27,K28,K29,K30,K31,K32,K33,K34,K35,K36,K37,K38,K40,K41,K42,K43,K44,K45,K46,K47,K48,K52,K53,K54,K55,K56,K57,K59,K60,K62,K64,K65,K66,K67,K68,K69,K71,K72,K75,K76,K77,K78,L02,K73,K74 |
| Major Vessel | L01,L04,L05,L06,L07,L08,L09,L10,L12,L13,L16,L18,L19,L20,L21,L22,L23,L25,L26,L27,L28,L29,L30,L31,L37,L38,L39,L41,L42,L43,L45,L46,L47,L48,L49,L50,L51,L52,L53,L54,L69,L77,L79,L80 |
| Vascular | L03,L56,L57,L58,L59,L60,L62,L63,L65,L66,L67,L68,L70,L71,L73,L74,L75,L76,L81,L82,L83,L84,L85,L86,L87,L88,L89,L90,L91,L93,L94,L96,L97,L98,L99,O01,O02,O03,O04,O15,X07,X08,X09,X10,X11,X12,O20 |
| Cerebrovascular | L33,L34,L35,O05 |
| Urological | M01,M02,M03,M04,M05,M06,M08,M09,M10,M13,M15,M16,M17,M18,M19,M20,M21,M22,M23,M25,M26,M27,M28,M29,M32,M33,M34,M35,M36,M37,M38,M39,M41,M42,M43,M44,M48,M49,M51,M52,M53,M54,M55,M56,M58,M60,M61,M62,M64,M65,M66,M67,M68,M70,M71,M72,M73,M75,M76,M79,M81,M83,M86,X15 |
| Male GU | N01,N03,N05,N06,N07,N08,N09,N10,N11,N13,N15,N17,N18,N19,N20,N22,N24,N26,N27,N28,N29,N30,N32,N34 |
| Orthopaedics | O06,O07,O08,O09,O10,O17,O18,O19,O21,O22,O23,O24,O25,O26,O27,O29 |
| Female LGU | P01,P03,P05,P06,P07,P09,P11,P13,P14,P15,P17,P18,P19,P20,P21,P22,P23,P24,P25,P29,P31,P32,M57,P28,P30 |
| Female UGU | Q01,Q02,Q05,Q07,Q08,Q09,Q10,Q11,Q16,Q17,Q19,Q20,Q22,Q23,Q24,Q25,Q26,Q27,Q28,Q29,Q30,Q31,Q32,Q34,Q35,Q36,Q37,Q38,Q39,Q41,Q43,Q44,Q45,Q47,Q49,Q50,Q51,Q52,Q54,Q56,R06,Q57 |
| Obstetrics | R01,R02,R04,R05,R07,R08,R10,R12,R17,R18,R28,R29,R30,R34 |
| Skin | S01,S02,S03,S04,S05,S06,S10,S11,S17,S18,S19,S20,S21,S22,S23,S24,S25,S26,S27,S28,S30,S31,S33,S34,S35,S36,S37,S38,S39,S40,S41,S42,S47,S48,S49,S54,S55,S56,S57,S60,S62,S64,S66,S68,S70,T59,T60,T61,T94 |
| Muscle | T50,T52,T53,T54,T55,T56,T57,T64,T65,T67,T68,T69,T70,T71,T72,T74,T76,T77,T79,T80,T83,W72,W73,W74,W75,W76 |
| Joint | T62,W01,W37,W38,W39,W40,W41,W42,W43,W44,W45,W46,W47,W48,W49,W50,W51,W55,W56,W57,W58,W59,W60,W61,W62,W63,W64,W65,W67,W69,W70,W71,W77,W78,W79,W80,W81,W82,W83,W84,W85,W86,W87,W88,W89,W91,W92,W93,W94,W95,W96,W97,W98 |
| Other | T85,T86,T87,T88,T89,T91,T92,T96,W34,W99,X03,X16,X46,X53,X55,X04,X17 |
| Skull & Spine | V01,V02,V03,V04,V05,V06,V07,V08,V09,V10,V11,V12,V13,V14,V15,V16,V17,V18,V19,V20,V21,V22,V23,V24,V25,V26,V27,V28,V29,V30,V31,V32,V33,V34,V35,V36,V37,V38,V39,V40,V41,V42,V43,V44,V45,V46,V47,V48,V49,V52,V54,V56,V58,V60,V62,V66,V67,V68,V57,V61,V51 |

| | |
|---|---|
| Bone | W02,W03,W04,W05,W06,W07,W08,W09,W10,W11,W12,W13,W14,W15,W16,W17,W18,W19,W20,W21,W22,W23,W24, W25,W26,W27,W28,W29,W30,W31,W32,W33,W52,W53,W54,W68,X01,X02,X05,X19,X20,X21,X22,X23,X24,X25,X27 |
| ORGAN DONO R | X45 |

**Appendix A OPCS4.7 codes defining surgical procedures, stratified by anatomical location of primary procedure.**

| Anatomical Location | OPCS 4.7 Codes – Class two procedures |
|---|---|
| Bone | W19,W20,W21,W22,W23,W24,W25,W26,W68,X01 |
| Breast | B28 |
| Cardiac | K54,K69 |
| Ear | D04 |
| Endocrine | B06,B20,B23,B25 |
| Female UGU | Q11,R06 |
| HPB | J06,J10,J11,J12,J13,J15,J16,J24,J25,J35,J52,J54,J56,J60,J61,J63,J65,J68,J72,J73 |
| Joint | W65,W67 |
| Lower GI | H06,H07,T36,T39,T45,T48,X14 |
| Major Vessel | L29,L48 |
| Male GU | N26 |
| Neuro | A02,A04,A08,A10,A41,A47,A62,A64 |
| ORGAN DONOR | X45 |
| Obstetrics | R01,R04,R05,R07,R08,R10,R12,R18,R28,R29,R30,R34 |
| Ocular | C08,C54,C55,C57,C69,C81,C85,C88 |
| Oral | F22,F24 |
| Orthopaedics | O17 |
| Other | T85,T86,T87,T88,T91,W99,X53,X55,X17 |
| Pharynx | E19,E29 |
| Skin | S21,S35,S40,S41,S42,S47,S54,S55,S56,S57,S66 |
| Skull & Spine | V08,V09,V15,V47 |
| Thoracic | E47,E50,E52,E54,E59,E63,T07,T08,T09,T10,T11,T12,T13,T14 |
| Upper GI | G01,G02,G03,G13,G27,G41,G68 |
| Urological | M13,M15,M17,M83 |
| Vascular | L56,L67,L68,L89,L91,L94,L96,L99,O15 |

**Appendix B (Class 2). OPCS4.7 codes defining class two procedures, stratified by anatomical location of primary procedure.** Class two procedures are elective operations that need to be performed within four weeks and were classified based on historical waiting times (from 2014-2019).

| Anatomical location | OPCS 4.7 Codes – Class three procedures |
|---|---|
| Bone | W05,W09,W10,W11,W18,W28,W29,W30,W31,W32,W33,X05 |
| Breast | B27,B30,B33,B34,B35,B40 |
| Cardiac | K02,K04,K07,K08,K09,K10,K11,K14,K15,K16,K20,K22,K23,K26,K30,K31,K32,K35,K37,K38,K40,K41,K42,K43,K44,K45,K46,K47,K48,K52,K55,K56,K59,K60,K65,K66,K67,K68,K71,K72,K75,K77,K78,L02,K73,K74 |
| Cerebrovascular | L33,L34,L35,O05 |
| Ear | D01,D02,D06,D08,D15,D19,D20,D23,D24,D26,D28 |
| Endocrine | B01,B04,B08,B09,B12,B17,B18,B22 |
| Female LGU | P01,P03,P05,P06,P07,P09,P11,P13,P15,P17,P19,P20,P29,P31,M57,P28,P30 |
| Female UGU | Q01,Q02,Q05,Q07,Q08,Q10,Q16,Q17,Q19,Q20,Q22,Q23,Q24,Q25,Q26,Q27,Q28,Q29,Q31,Q32,Q34,Q35,Q36,Q37,Q38,Q39,Q41,Q43,Q44,Q45,Q47,Q49,Q50,Q51,Q52,Q57 |
| HPB | J02,J03,J04,J05,J07,J08,J18,J20,J21,J23,J26,J27,J28,J29,J30,J31,J32,J33,J34,J36,J37,J49,J55,J57,J58,J59,J62,J69,J70,J77 |
| Joint | T62,W49,W69,W70,W78,W81,W82,W83,W85,W86,W87,W89,W91,W92 |
| Lower GI | H01,H02,H04,H05,H08,H09,H10,H11,H12,H13,H14,H15,H16,H17,H19,H29,H30,H33,H40,H41,H44,H46,H48,H49,H51,H52,H53,H54,H55,H56,H58,H59,H60,H62,H66,T19,T20,T21,T22,T23,T24,T27,T29,T30,T31,T33,T34,T38,T41,T42,T43,H37 |
| Major Vessel | L01,L06,L07,L08,L09,L10,L12,L16,L18,L19,L20,L21,L22,L23,L25,L26,L27,L28,L30,L31,L37,L38,L39,L41,L42,L43,L45,L46,L47,L49,L50,L51,L52,L53,L54,L69,L79,L80 |
| Male GU | N01,N03,N05,N06,N07,N13,N17,N18,N19,N20,N22,N24,N27,N30,N32 |
| Muscle | T53,T54,T57,T65,T67,T68,T71,T72,T74,T77,T79,T83,W75,W76 |
| Nasal | E01,E05,E08,E09,E10,E11,E12,E16,E17,E64,E66 |
| Neuro | A01,A05,A06,A09,A11,A12,A13,A14,A16,A17,A18,A20,A22,A26,A27,A28,A29,A30,A31,A33,A34,A38,A39,A40,A42,A43,A44,A45,A48,A49,A59,A61,A63,A65,A67,A68,A69,A75,A76,A77,A84,A82 |
| Obstetrics | R17 |
| Ocular | C01,C02,C03,C06,C10,C11,C12,C14,C15,C16,C17,C19,C20,C22,C24,C26,C27,C29,C39,C40,C41,C43,C45,C47,C49,C51,C52,C53,C59,C60,C61,C62,C64,C65,C66,C67,C71,C73,C74,C75,C77,C79,C80,C82,C84,C86,C89 |
| Oral | F01,F02,F04,F05,F06,F09,F18,F23,F26,F28,F32,F36,F38,F39,F40,F42,F44,F45,F46,F48,F51,F53,F58 |
| Orthopaedics | O10,O19,O23,O24,O25,O26 |
| Other | T92,T96,W34,X03,X46 |
| Pharynx | E23,E24,E27,E30,E31,E33,E34,E35,E38 |
| Skin | S04,S06,S10,S11,S17,S22,S24,S25,S26,S27,S28,S30,S34,S36,S37,S38,S49,S64,S68,S70,T59,T60,T61,T94 |
| Skull & Spine | V01,V03,V04,V05,V06,V07,V11,V14,V17,V18,V19,V22,V24,V30,V31,V33,V35,V43,V44,V45,V56,V57 |
| Thoracic | E39,E40,E41,E42,E43,E44,E46,E48,E55,E57,E61,E62,T01,T03,T05,E67 |
| Upper GI | G04,G05,G06,G07,G08,G10,G11,G14,G17,G21,G29,G30,G34,G35,G36,G38,G40,G48,G49,G50,G51,G52,G53,G57,G58,G59,G60,G61,G63,G67,G69,G70,G71,G73,G74,G75,G76,G78,G82,T15,T16,T17,T37,G12,G20 |
| Urological | M01,M02,M03,M04,M05,M06,M08,M09,M10,M16,M18,M21,M22,M23,M25,M26,M27,M28,M29,M32,M33,M34,M35,M37,M38,M41,M42,M44,M49,M58,M60,M61,M62,M67,M68,M70,M71,M75,M76,M79,M81,M86 |
| Vascular | L03,L57,L58,L59,L60,L62,L63,L65,L66,L71,L73,L74,L75,L76,L81,L82,L90,L93,L97,O01,O02,O03,O04,X07,X08,X09,X10,X11,X12 |

**Appendix B (Class 3). OPCS4.7 codes defining class three procedures, stratified by anatomical location of primary procedure.** Class three procedures are elective operations that need to be performed within three months and were classified based on historical waiting times (from 2014-2019).

| Anatomical Location | OPCS 4.7 Codes – Class four procedures |
|---|---|
| Bone | W02,W03,W04,W06,W07,W08,W12,W13,W14,W15,W16,W17,W27,W52,W53,W54,X02,X19,X20,X21,X22,X23,X24,X25,X27 |
| Breast | B29,B31,B36,B37,B38,B39 |
| Cardiac | K01,K05,K06,K12,K13,K17,K18,K19,K24,K25,K27,K28,K29,K33,K34,K36,K53,K57,K62,K64,K76 |
| Ear | D03,D10,D12,D13,D14,D16,D17,D22 |
| Endocrine | B02,B10,B14,B16 |
| Female LGU | P14,P18,P21,P22,P23,P24,P25,P32 |
| Female UGU | Q09,Q30,Q54,Q56 |
| HPB | J01,J19 |
| Joint | W01,W37,W38,W39,W40,W41,W42,W43,W44,W45,W46,W47,W48,W50,W51,W55,W56,W57,W58,W59,W60,W61,W62,W63,W64,W71,W77,W79,W80,W84,W88,W93,W94,W95,W96,W97,W98 |
| Lower GI | H03,H32,H34,H35,H36,H42,H47,H50,H57,T25,T26,T28,T51,T97,T98,T32 |
| Major Vessel | L04,L05,L13,L77 |
| Male GU | N08,N09,N10,N11,N15,N28,N29,N34 |
| Muscle | T50,T52,T55,T56,T64,T69,T70,T76,T80,W72,W73,W74 |
| Nasal | E02,E03,E04,E07,E13,E14,E15 |
| Neuro | A03,A07,A24,A25,A32,A36,A51,A57,A60,A66,A70,A73,A78,A79,A81 |
| Obstetrics | R02 |
| Ocular | C05,C09,C13,C18,C23,C25,C31,C32,C33,C34,C35,C37,C44,C46,C72,C83 |
| Oral | F03,F11,F29,F30,F50,F52 |
| Orthopaedics | O06,O07,O08,O09,O18,O21,O22,O27,O29 |
| Other | T89,X16,X04 |
| Pharynx | E20,E21,E28,F34 |
| Skin | S01,S02,S03,S05,S18,S19,S20,S23,S31,S33,S39,S48,S60,S62 |
| Skull & Spine | V02,V10,V12,V13,V16,V20,V21,V23,V25,V26,V27,V28,V29,V32,V34,V36,V37,V38,V39,V40,V41,V42,V46,V48,V49,V52,V54,V58,V60,V62,V66,V67,V68,V61,V51 |
| Thoracic | E53,T02 |
| Upper GI | G09,G23,G24,G25,G26,G28,G31,G32,G33,G72 |
| Urological | M19,M20,M36,M39,M43,M48,M51,M52,M53,M54,M55,M56,M64,M65,M66,M72,M73,X15 |
| Vascular | L70,L83,L84,L85,L86,L87,L88,L98,O20 |

**Appendix B (Class 4). OPCS4.7 codes defining class four procedures, stratified by anatomical location of primary procedure.** Class four procedures are elective operations that can be delayed beyond four months and were classified based on historical waiting times (from 2014-2019).

**Appendix C Flow diagram demonstrating number of patients identified in study cohort**

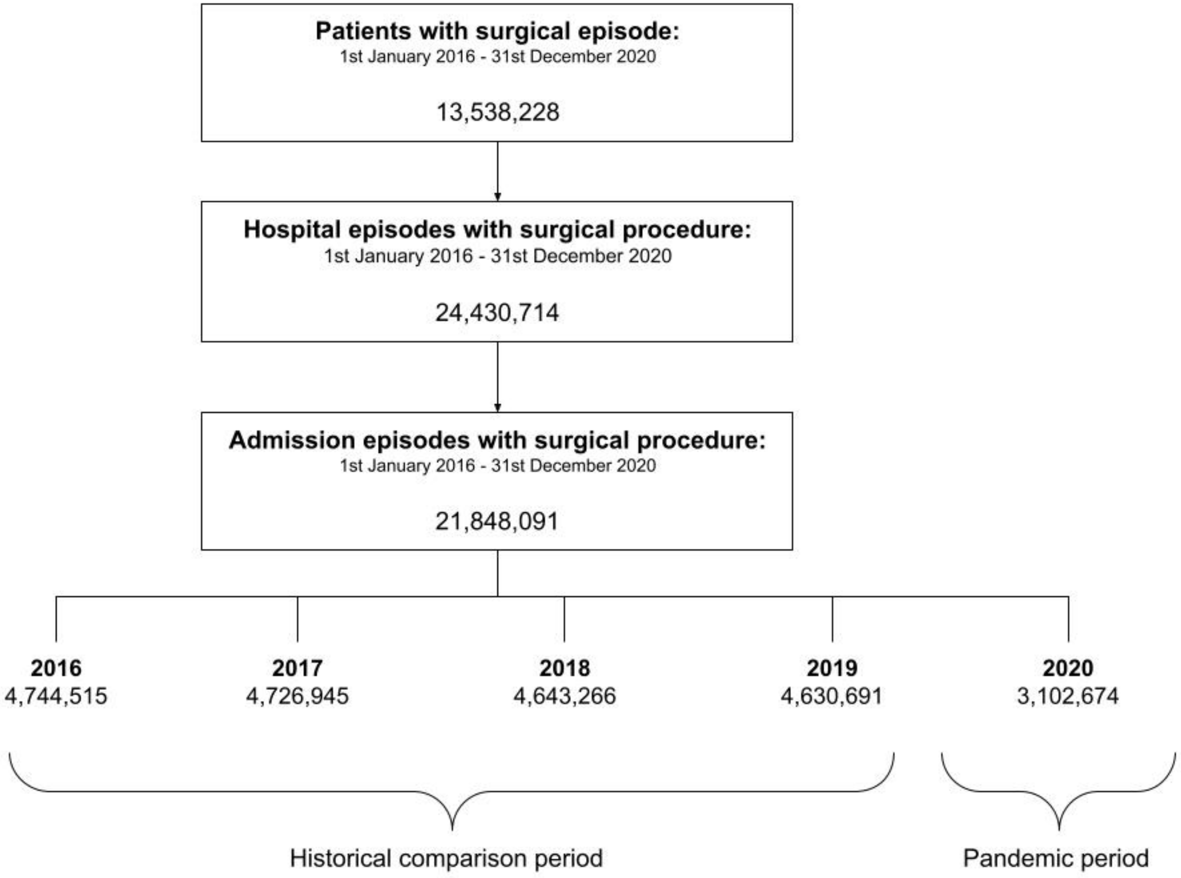

**Appendix D Wales/England Breakdown**

**Class Breakdown for England**

| | Class 1 | | Class 2 | | Class 3 | | Class 4 | |
|---|---|---|---|---|---|---|---|---|
| | Observed, N | Percentage of expected (95%CI) | Observed | Percentage of expected (95%CI) | Observed | Percentage of expected (95%CI) | Observed | Percentage of expected (95%CI) |
| January | 68,597 | 99.6 (92.4-107.9) | 33,984 | 106.0 (95.6-119.2) | 206,058 | 101.3 (91.5-113.4) | 71,976 | 98.1 (84.2-117.7) |
| February | 63,620 | 92.3 (85.7-100.1) | 30,853 | 96.3 (86.7-108.2) | 191,939 | 97.5 (87.8-109.6) | 68,825 | 89.9 (77.6-107.0) |
| March | 59,271 | 86.0 (79.9-93.2) | 28,724 | 89.6 (80.7-100.7) | 143,061 | 69.6 (63.0-77.9) | 42,676 | 58.2 (49.9-69.8) |
| April | 46,411 | 67.4 (62.5-73.0) | 16,125 | 50.3 (45.3-65.5) | 35,452 | 18.4 (16.5-20.7) | 1,570 | 2.3 (1.9-2.8) |
| May | 56,695 | 82.3 (76.4-89.2) | 17,618 | 54.9 (49.5-61.7) | 45,157 | 22.9 (20.7-25.8) | 3,441 | 4.6 (3.9-5.6) |
| June | 61,054 | 88.6 (82.3-76.4) | 21,249 | 66.2 (59.6-74.4) | 74,744 | 38.4 (34.6-43.2) | 10,741 | 14.4 (12.2-17.6) |
| July | 67,040 | 97.3 (90.3-105.5) | 24,325 | 75.8 (68.2-85.2) | 109,011 | 53.7 (48.5-60.1) | 23,129 | 31.0 (26.2-37.9) |
| August | 65,334 | 94.8 (88.0-102.8) | 22,524 | 70.1 (63.1-78.8) | 114,872 | 60.0 (53.8-67.6) | 31,273 | 41.9 (35.5-51.3) |
| September | 65,265 | 94.7 (87.9-102.7) | 26,377 | 82.1 (73.9-92.3) | 150,609 | 76.1 (68.6-85.5) | 45,413 | 60.9 (51.5-74.5) |
| October | 64,084 | 93.0 (86.3-100.8) | 27,179 | 84.5 (76.1-95.1) | 162,505 | 80.6 (72.8-90.3) | 52,628 | 70.6 (59.7-86.3) |
| November | 59,246 | 86.0 (79.8-93.2) | 26,279 | 81.7 (73.6-91.9) | 158,763 | 80.4 (72.4-90.4) | 49,159 | 65.9 (55.8-80.6) |
| December | 49,077 | 71.2 (66.1-77.2) | 23,721 | 73.7 (66.4-82.9) | 129,078 | 68.6 (61.5-77.6) | 34,429 | 46.2 (39.1-56.4) |

**Class breakdown for Wales**

| | Class 1 | | Class 2 | | Class 3 | | Class 4 | |
|---|---|---|---|---|---|---|---|---|
| | Observed, N | Percentage of expected (95%CI) | Observed | Percentage of expected (95%CI) | | Percentage of expected (95%CI) | Observed | Percentage of expected (95%CI) |
| January | 3,931 | 103.0 (95.8-112.0) | 1,083 | 104.0 (89.1-124.9) | 10,397 | 107.5 (92.3-128.80 | 3,219 | 92.0 (76.3-115.9) |
| February | 3,521 | 93.8 (86.9-101.8) | 986 | 94.7 (81.2-113.7) | 9,724 | 100.6 (86.3-120.5) | 3,518 | 100.5 (83.4-126.6) |
| March | 3,084 | 79.5 (73.9-86.0) | 905 | 86.9 (74.5-104.4) | 6,681 | 69.1 (59.3-82.8) | 1,820 | 52.0 (43.1-65.6) |
| April | 2,387 | 61.3 (57.0-66.3) | 400 | 38.4 (32.9-46.1) | 1,668 | 17.3 (14.8-20.7) | 50 | 1.4 (1.2-1.8) |
| May | 2,902 | 75.4 (70.0-81.6) | 394 | 37.8 (32.4-45.4) | 2,117 | 21.9 (18.8-26.2) | 101 | 2.9 (2.4-3.6) |
| June | 3,124 | 84.8 (78.5-92.2) | 523 | 50.2 (43.0-60.3) | 3,231 | 33.4 (28.7-40.0) | 292 | 8.3 (6.9-10.5) |
| July | 3,593 | 95.3 (88.4-103.4) | 663 | 63.7 (54.6-76.5) | 4,130 | 42.7 (36.7-51.2) | 490 | 14.0 (11.6-17.6) |
| August | 3,564 | 92.2 (85.6-99.8) | 659 | 63.3 (54.2-76.0) | 3,876 | 40.1 (34.4-48.0) | 512 | 14.6 (12.1-18.4) |
| September | 3,677 | 96.6 (89.7-104.8) | 768 | 73.8 (63.2-88.6) | 4,960 | 51.3 (44.0-61.5) | 826 | 23.6 (19.6-29.7) |
| October | 3,230 | 86.0 (79.7-93.4) | 754 | 72.4 (62.1-87.0) | 5,521 | 57.1 (49.0-68.4) | 890 | 25.4 (21.1-32.0) |
| November | 2,879 | 75.6 (70.2-82.0) | 728 | 69.9 (59.9-84.0) | 5,297 | 54.8 (47.0-65.6) | 995 | 28.4 (23.6-35.8) |
| December | 2,144 | 56.8 (52.7-61.6) | 592 | 56.9 (48.7-68.3) | 3,957 | 40.9 (35.1-49.0) | 750 | 21.4 (17.8-27.0) |

### Appendix E Anatomical Breakdown

| | Month | January | February | March | April | May | June | July | August | September | October | November | December |
|---|---|---|---|---|---|---|---|---|---|---|---|---|---|
| **Bone** | **Observed** | 18,099 | 16,533 | 13,909 | 6,784 | 9,253 | 11,184 | 14,048 | 14,669 | 16,126 | 15,384 | 13,316 | 10,207 |
| | **Expected** | 17,178 | 17,178 | 17,178 | 17,178 | 17,178 | 17,178 | 17,178 | 17,178 | 17,178 | 17,178 | 17,178 | 17,178 |
| | **95% CI** | 14844-19513 | 14487-19869 | 14173-20184 | 13888-20468 | 13626-20730 | 13382-20974 | 13153-21203 | 12936-21420 | 12730-21626 | 12533-21823 | 12344-22012 | 12162-22194 |
| **Breast** | **Observed** | 7,513 | 6,864 | 6,445 | 4,210 | 3,493 | 3,766 | 4,481 | 4,496 | 5,777 | 6,045 | 6,118 | 5,261 |
| | **Expected** | 7,726 | 7,305 | 7,556 | 7,348 | 7,496 | 7,355 | 7,580 | 7,195 | 7,389 | 7,454 | 7,333 | 7,021 |
| | **95% CI** | 6786-8665 | 6365-8244 | 6616-8496 | 6408-8288 | 6557-8436 | 6415-8295 | 6640-8520 | 6255-8135 | 6450-8329 | 6515-8394 | 6393-8272 | 6081-7961 |
| **Cardiac** | **Observed** | 13,834 | 12,915 | 11,720 | 6,330 | 7,982 | 10,668 | 12,530 | 11,156 | 12,091 | 12,243 | 11,567 | 9,509 |
| | **Expected** | 13,427 | 13,427 | 13,427 | 13,427 | 13,427 | 13,427 | 13,427 | 13,427 | 13,427 | 13,427 | 13,427 | 13,427 |
| | **95% CI** | 11999-14854 | 11999-14854 | 11999-14854 | 11999-14854 | 11999-14854 | 11999-14854 | 11999-14854 | 11999-14854 | 11999-14854 | 11999-14854 | 11999-14854 | 11999-14854 |
| **Cerebrovascular** | **Observed** | 432 | 432 | 294 | 150 | 256 | 359 | 428 | 489 | 516 | 494 | 493 | 388 |
| | **Expected** | 399 | 399 | 399 | 400 | 400 | 400 | 400 | 401 | 401 | 401 | 401 | 402 |
| | **95% CI** | 332-465 | 333-466 | 333-466 | 333-466 | 333-467 | 334-467 | 334-467 | 334-467 | 334-468 | 335-468 | 335-468 | 335-468 |
| **Ear** | **Observed** | 6,474 | 5,966 | 3,890 | 741 | 1,047 | 1,692 | 2,498 | 2,917 | 3,895 | 4,314 | 4,202 | 3,236 |
| | **Expected** | 6,182 | 6,159 | 6,136 | 6,113 | 6,090 | 6,067 | 6,044 | 6,021 | 5,998 | 5,975 | 5,952 | 5,929 |
| | **95% CI** | 5281-7082 | 5243-7074 | 5205-7067 | 5165-7061 | 5124-7056 | 5082-7052 | 5039-7049 | 4995-7047 | 4949-7047 | 4903-7047 | 4856-7049 | 4808-7051 |
| **Endocrine** | **Observed** | 1,968 | 1,850 | 1,379 | 339 | 634 | 1,111 | 1,519 | 1,333 | 1,626 | 1,741 | 1,640 | 1,236 |
| | **Expected** | 1,963 | 1,870 | 1,955 | 1,883 | 1,881 | 1,863 | 1,946 | 1,798 | 1,932 | 1,900 | 1,898 | 1,781 |
| | **95% CI** | 1693-2233 | 1600-2140 | 1685-2225 | 1613-2153 | 1611-2151 | 1593-2133 | 1676-2216 | 1528-2068 | 1662-2202 | 1630-2170 | 1628-2168 | 1511-2051 |
| **Female LGU** | **Observed** | 3,906 | 3,692 | 2,607 | 554 | 785 | 1,242 | 1,760 | 2,119 | 2,600 | 2,912 | 2,761 | 1,957 |
| | **Expected** | 3,753 | 3,731 | 3,709 | 3,687 | 3,665 | 3,643 | 3,621 | 3,599 | 3,577 | 3,555 | 3,533 | 3,511 |
| | **95% CI** | 3018-4488 | 2994-4468 | 2970-4449 | 2945-4429 | 2921-4409 | 2896-4390 | 2872-4370 | 2848-4350 | 2823-4331 | 2799-4311 | 2775-4291 | 2750-4271 |
| **Female UGU** | **Observed** | 16,442 | 15,105 | 12,271 | 4,965 | 5,838 | 7,505 | 9,546 | 9,969 | 12,232 | 13,254 | 12,578 | 10,297 |
| | **Expected** | 16,925 | 16,442 | 17,449 | 16,106 | 16,438 | 16,180 | 16,809 | 15,895 | 16,329 | 16,717 | 16,082 | 15,411 |
| | **95% CI** | 15092-18758 | 14609-18275 | 15616-19282 | 14273-17939 | 14605-18271 | 14347-18013 | 14976-18642 | 14062-17728 | 14496-18162 | 14884-18550 | 14249-17915 | 13578-17244 |
| **HPB** | **Observed** | 8,466 | 7,827 | 5,752 | 1,598 | 2,580 | 4,390 | 6,214 | 6,208 | 7,419 | 7,577 | 7,342 | 5,381 |
| | **Expected** | 8,516 | 8,371 | 8,534 | 8,184 | 8,332 | 8,269 | 8,516 | 8,108 | 8,253 | 8,376 | 8,178 | 7,970 |

| | | | | | | | | | | | | | |
|---|---|---|---|---|---|---|---|---|---|---|---|---|---|
| | **95% CI** | 7393-9639 | 7248-9493 | 7411-9657 | 7062-9307 | 7210-9455 | 7146-9391 | 7393-9639 | 6985-9231 | 7130-9375 | 7253-9499 | 7055-9300 | 6847-9093 |
| **Joint** | **Observed** | 31,056 | 30,121 | 19,992 | 3,608 | 4,549 | 6,469 | 10,888 | 14,391 | 19,890 | 22,025 | 19,731 | 13,799 |
| | **Expected** | 29,918 | 29,797 | 29,675 | 29,554 | 29,432 | 29,311 | 29,190 | 29,068 | 28,947 | 28,825 | 28,704 | 28,582 |
| | **95% CI** | 25191-34645 | 25059-34534 | 24928-34423 | 24796-34312 | 24664-34201 | 24533-34089 | 24401-33978 | 24270-33867 | 24138-33755 | 24006-33644 | 23875-33533 | 23743-33421 |
| **Lower GI** | **Observed** | 31,095 | 28,874 | 21,272 | 6,961 | 9,737 | 13,788 | 18,547 | 19,824 | 23,745 | 25,259 | 23,967 | 18,684 |
| | **Expected** | 32,077 | 30,853 | 32,478 | 30,551 | 31,334 | 30,728 | 31,577 | 30,174 | 30,777 | 31,444 | 30,728 | 29,656 |
| | **95% CI** | 28890-35264 | 27666-34040 | 29291-35665 | 27364-33737 | 28147-34521 | 27541-33915 | 28390-34764 | 26987-33361 | 27590-33963 | 28257-34631 | 27541-33915 | 26469-32842 |
| **Major Vessel** | **Observed** | 1,906 | 1,793 | 1,459 | 639 | 878 | 1,323 | 1,629 | 1,429 | 1,652 | 1,637 | 1,567 | 1,216 |
| | **Expected** | 1,833 | 1,826 | 1,820 | 1,813 | 1,806 | 1,800 | 1,793 | 1,786 | 1,779 | 1,773 | 1,766 | 1,759 |
| | **95% CI** | 1559-2107 | 1551-2102 | 1544-2096 | 1536-2090 | 1528-2084 | 1521-2078 | 1513-2072 | 1506-2067 | 1498-2061 | 1491-2055 | 1483-2049 | 1476-2043 |
| **Male GU** | **Observed** | 7,208 | 6,805 | 4,726 | 1,116 | 1,375 | 2,011 | 3,153 | 3,649 | 4,871 | 5,587 | 5,226 | 4,289 |
| | **Expected** | 7,264 | 7,034 | 7,415 | 6,852 | 7,070 | 6,957 | 7,192 | 6,922 | 7,101 | 7,256 | 6,940 | 6,613 |
| | **95% CI** | 6369-8159 | 6138-7929 | 6520-8311 | 5957-7748 | 6175-7966 | 6061-7852 | 6296-8087 | 6026-7817 | 6206-7997 | 6360-8151 | 6044-7835 | 5718-7508 |
| **Muscle** | **Observed** | 9,148 | 8,328 | 5,769 | 1,067 | 1,619 | 2,809 | 4,187 | 4,618 | 5,950 | 6,319 | 5,901 | 4,962 |
| | **Expected** | 8,806 | 9,190 | 8,779 | 8,510 | 8,923 | 8,923 | 8,923 | 8,923 | 8,923 | 8,923 | 8,923 | 8,923 |
| | **95% CI** | 7698-9915 | 8082-10299 | 7670-9887 | 7401-9618 | 7719-10126 | 7719-10126 | 7719-10126 | 7719-10126 | 7719-10126 | 7719-10126 | 7719-10126 | 7719-10126 |
| **Nasal** | **Observed** | 7,822 | 7,404 | 5,081 | 1,041 | 1,665 | 2,675 | 3,566 | 3,795 | 4,997 | 5,637 | 5,397 | 4,084 |
| | **Expected** | 7,565 | 7,565 | 7,565 | 7,565 | 7,565 | 7,565 | 7,565 | 7,565 | 7,565 | 7,565 | 7,565 | 7,565 |
| | **95% CI** | 6513-8617 | 6513-8617 | 6513-8617 | 6513-8617 | 6513-8617 | 6513-8617 | 6513-8617 | 6513-8617 | 6513-8617 | 6513-8617 | 6513-8617 | 6513-8617 |
| **Neuro** | **Observed** | 19,342 | 17,838 | 11,984 | 2,046 | 3,019 | 5,057 | 8,288 | 9,208 | 12,245 | 13,784 | 13,282 | 10,603 |
| | **Expected** | 18,244 | 18,483 | 18,201 | 17,402 | 17,888 | 17,888 | 17,888 | 17,888 | 17,888 | 17,888 | 17,888 | 17,888 |
| | **95% CI** | 16108-20380 | 16245-20722 | 15963-20440 | 15163-19640 | 15550-20226 | 15550-20226 | 15550-20226 | 15550-20226 | 15550-20226 | 15550-20226 | 15550-20226 | 15550-20226 |
| **Obstetrics** | **Observed** | 16,031 | 14,813 | 15,597 | 14,935 | 15,581 | 15,967 | 16,777 | 15,840 | 16,713 | 16,808 | 15,585 | 13,511 |
| | **Expected** | 15,652 | 15,652 | 15,652 | 15,652 | 15,652 | 15,652 | 15,652 | 15,652 | 15,652 | 15,652 | 15,652 | 15,652 |
| | **95% CI** | 14291-17013 | 14291-17013 | 14291-17013 | 14291-17013 | 14291-17013 | 14291-17013 | 14291-17013 | 14291-17013 | 14291-17013 | 14291-17013 | 14291-17013 | 14291-17013 |
| **Ocular** | **Observed** | 70,970 | 65,558 | 47,786 | 8,490 | 10,763 | 19,726 | 33,502 | 37,947 | 51,813 | 56,198 | 56,224 | 46,659 |
| | **Expected** | 65,061 | 62,895 | 66,871 | 61,458 | 63,456 | 62,630 | 66,063 | 61,951 | 64,748 | 65,774 | 64,754 | 61,013 |
| | **95% CI** | 57276-72845 | 55110-70679 | 59087-74656 | 53674-69243 | 55672-71240 | 54846-70414 | 58278-73847 | 54167-69736 | 56964-72532 | 57990-73559 | 56970-72538 | 53229-68797 |
| **Oral** | **Observed** | 15,114 | 14,463 | 9,168 | 1,455 | 2,228 | 3,760 | 5,677 | 6,337 | 8,285 | 10,068 | 9,876 | 7,139 |

| | | | | | | | | | | | | | |
|---|---|---|---|---|---|---|---|---|---|---|---|---|---|
| | **Expected** | 15,602 | 15,043 | 15,646 | 14,704 | 14,786 | 14,853 | 15,450 | 14,413 | 14,694 | 15,098 | 14,648 | 13,860 |
| | **95% CI** | 13502-17702 | 12943-17143 | 13546-17746 | 12604-16804 | 12686-16886 | 12753-16952 | 13350-17550 | 12313-16512 | 12594-16794 | 12998-17198 | 12548-16748 | 11760-15960 |
| **Organ donor** | **Observed** | 111 | 80 | 38 | 20 | 20 | 37 | 41 | 38 | 67 | 59 | 57 | 52 |
| | **Expected** | 77 | 77 | 77 | 77 | 77 | 77 | 77 | 77 | 77 | 77 | 77 | 77 |
| | **95% CI** | 60-95 | 60-95 | 60-95 | 60-95 | 60-95 | 60-95 | 60-95 | 60-95 | 60-95 | 60-95 | 60-95 | 60-95 |
| **Orthopaedics** | **Observed** | 2,267 | 2,170 | 1,464 | 131 | 217 | 569 | 1,023 | 1,157 | 1,575 | 1,577 | 1,477 | 1,096 |
| | **Expected** | 2,324 | 2,208 | 2,156 | 2,008 | 2,171 | 2,042 | 2,040 | 1,958 | 2,020 | 1,922 | 1,929 | 1,868 |
| | **95% CI** | 1898-2750 | 1781-2635 | 1709-2603 | 1554-2462 | 1649-2693 | 1520-2564 | 1492-2588 | 1402-2514 | 1434-2607 | 1332-2513 | 1316-2542 | 1246-2489 |
| **Other** | **Observed** | 2,552 | 2,230 | 2,087 | 1,409 | 1,432 | 1,622 | 1,887 | 1,660 | 2,106 | 2,123 | 2,108 | 1,898 |
| | **Expected** | 2,324 | 2,317 | 2,311 | 2,304 | 2,298 | 2,292 | 2,285 | 2,279 | 2,272 | 2,266 | 2,259 | 2,253 |
| | **95% CI** | 2051-2596 | 2045-2590 | 2038-2584 | 2031-2578 | 2024-2572 | 2017-2566 | 2010-2560 | 2003-2554 | 1997-2548 | 1990-2542 | 1983-2536 | 1976-2530 |
| **Pharynx** | **Observed** | 5,387 | 5,295 | 3,171 | 353 | 470 | 1,016 | 1,873 | 2,514 | 3,252 | 3,653 | 3,435 | 2,617 |
| | **Expected** | 5,211 | 5,191 | 5,172 | 5,153 | 5,134 | 5,115 | 5,096 | 5,077 | 5,058 | 5,039 | 5,020 | 5,001 |
| | **95% CI** | 4194-6227 | 4172-6211 | 4149-6196 | 4127-6180 | 4105-6164 | 4083-6148 | 4061-6132 | 4038-6116 | 4016-6100 | 3994-6084 | 3972-6068 | 3950-6052 |
| **Skin** | **Observed** | 31,465 | 29,770 | 24,420 | 11,020 | 14,685 | 18,620 | 21,329 | 21,033 | 24,219 | 25,822 | 25,115 | 20,848 |
| | **Expected** | 31,333 | 30,638 | 31,912 | 30,471 | 30,752 | 30,506 | 32,006 | 30,685 | 31,022 | 31,679 | 30,932 | 29,767 |
| | **95% CI** | 28555-34110 | 27861-33416 | 29134-34689 | 27694-33249 | 27974-33529 | 27729-33284 | 29229-34784 | 27908-33463 | 28245-33800 | 28901-34456 | 28154-33709 | 26990-32545 |
| **Skull & Spine** | **Observed** | 10,014 | 9,896 | 6,496 | 974 | 1,483 | 2,790 | 4,410 | 5,062 | 6,804 | 7,219 | 7,128 | 5,261 |
| | **Expected** | 10,000 | 9,908 | 9,816 | 9,724 | 9,632 | 9,540 | 9,448 | 9,357 | 9,265 | 9,173 | 9,081 | 8,989 |
| | **95% CI** | 8237-11762 | 8137-11678 | 8037-11595 | 7937-11511 | 7837-11427 | 7738-11343 | 7638-11259 | 7538-11175 | 7438-11091 | 7339-11007 | 7239-10923 | 7140-10839 |
| **Thoracic** | **Observed** | 7,532 | 6,881 | 6,025 | 4,242 | 4,670 | 5,555 | 6,049 | 5,567 | 6,149 | 6,178 | 6,076 | 4,944 |
| | **Expected** | 7,358 | 7,071 | 7,343 | 7,026 | 7,058 | 7,016 | 7,502 | 7,147 | 7,200 | 7,291 | 7,121 | 7,011 |
| | **95% CI** | 6735-7982 | 6448-7694 | 6719-7966 | 6403-7649 | 6434-7681 | 6393-7639 | 6879-8126 | 6523-7770 | 6577-7823 | 6668-7915 | 6498-7745 | 6388-7634 |
| **Upper Gi** | **Observed** | 4,576 | 4,416 | 3,262 | 1,475 | 1,813 | 2,349 | 3,217 | 3,201 | 3,836 | 3,856 | 3,572 | 2,783 |
| | **Expected** | 4,663 | 4,666 | 4,669 | 4,672 | 4,675 | 4,678 | 4,681 | 4,684 | 4,687 | 4,690 | 4,692 | 4,695 |
| | **95% CI** | 4011-5315 | 4014-5318 | 4017-5321 | 4020-5324 | 4022-5327 | 4025-5330 | 4028-5333 | 4031-5336 | 4034-5339 | 4037-5342 | 4040-5345 | 4043-5348 |
| **Urological** | **Observed** | 29,182 | 27,256 | 22,633 | 8,583 | 10,798 | 15,313 | 19,998 | 19,397 | 23,077 | 23,605 | 23,250 | 19,531 |
| | **Expected** | 29,064 | 28,153 | 29,015 | 27,544 | 28,031 | 27,376 | 28,945 | 27,325 | 28,093 | 28,465 | 27,895 | 27,185 |
| | **95% CI** | 26261-31867 | 25350-30956 | 26212-31818 | 24741-30347 | 25228-30834 | 24573-30179 | 26142-31748 | 24522-30128 | 25290-30896 | 25662-31269 | 25092-30699 | 24382-29988 |

| | | | | | | | | | | | | | |
|---|---|---|---|---|---|---|---|---|---|---|---|---|---|
| Vascular | **Observed** | 19,388 | 17,859 | 15,573 | 8,864 | 9,620 | 11,640 | 13,378 | 12,658 | 14,419 | 15,465 | 14,399 | 12,334 |
| | **Expected** | 18,014 | 17,793 | 17,793 | 17,793 | 17,793 | 17,793 | 17,793 | 17,793 | 17,793 | 17,793 | 17,793 | 17,793 |
| | **95% CI** | 16267-19761 | 15885-19701 | 15885-19701 | 15885-19701 | 15885-19701 | 15885-19701 | 15885-19701 | 15885-19701 | 15885-19701 | 15885-19701 | 15885-19701 | 15885-19701 |
